# Associations between genetically predicted iron status and cardiovascular disease risk: A Mendelian randomization study

**DOI:** 10.1101/2024.02.05.24302373

**Authors:** Alexa Barad, Andrew G. Clark, Kimberly O. O’Brien, Eva K. Pressman

## Abstract

**Background:** Mendelian randomization (MR) studies suggest a causal effect of iron (Fe) status on cardiovascular disease (CVD) risk, but it is unknown if these associations are confounded by pleiotropic effects of the instrumental variables (IV) on CVD risk factors. We aimed to investigate the effect of Fe status on CVD risk controlling for CVD risk factors.

**Methods:** Fe biomarker IVs (total Fe binding capacity (TIBC, *n*=208,422), transferrin saturation (TSAT, *n*=198,516), serum Fe (SI, *n*=236,612), ferritin (*n*=257,953)) were selected from a European GWAS meta-analysis. We performed two-sample univariate (UV) MR of each Fe trait on CVD outcomes (all-cause ischemic stroke (IS), cardioembolic IS (CES), large artery IS (LAS), small vessel IS (SVS), and coronary heart disease (CHD)) from MEGASTROKE (*n*=440,328) and CARDIoGRAMplusC4D (*n*=183,305). We then implemented multivariate (MV) MR conditioning on six CVD risk factors from independent European samples to evaluate their potential confounding and/or mediating effects on the observed Fe-CVD associations.

**Results:** With UVMR analyses, we found higher genetically predicted Fe status to be associated with a greater risk of CES (TSAT: OR 1.17 [95%CI 1.03, 1.33], SI: OR 1.21 [ 95%CI 1.02, 1.44]; TIBC: OR 0.81 [95%CI 0.69, 0.94]). The detrimental effects of Fe status on CES risk remained unaffected when adjusting for CVD risk factors (all *P*<0.05). Additionally, we found diastolic blood pressure (DBP) to mediate between 7.1-8.8% of the total effect of Fe status on CES incidence. While UVMR initially suggested a protective effect of Fe status on LAS and CHD, MVMR analyses factoring CVD risk factors revealed a complete annulment of this perceived protective effect (all *P*>0.05).

**Discussion:** Higher Fe status was associated with a greater risk of CES independent of CVD risk factors, and this effect was partly mediated by DBP. These findings support a role of Fe status as a modifiable risk factor for CES.

## Introduction

Iron (Fe) is an essential nutrient needed to support many biological processes. Both extremes of Fe status have been associated with adverse cardiovascular outcomes. Iron deficiency is the most prevalent micronutrient deficiency worldwide and is associated with significant comorbidities affecting 70% of patients with heart failure (1), whereas Fe overload is implicated in numerous cardiometabolic diseases (2). When circulating Fe exceeds the transport carrying capacity of transferrin, Fe begins to circulate free, generating a toxic Fe species known as non-transferrin bound Fe (NTBI). Cardiac, pancreatic, and hepatic cells all internalize NTBI via different mechanisms (3, 4). Cellular uptake of NTBI increases the intracellular labile Fe pool resulting in generation of reactive oxygen species and subsequent oxidative tissue damage (5). Excess Fe in cardiomyocytes has been shown to induce ferroptosis, a form of regulated cell death driven by Fe-dependent lipid peroxidation that is linked to cardiovascular disease (CVD) (6).

Although a link between Fe overload and CVD risk was proposed over 40 years ago (7), epidemiological data to date have shown conflicting results. Clinical studies have reported associations between atherosclerosis and increased serum Fe (8) or serum ferritin (9–13) concentrations. Consistent with these observations, some studies demonstrated an association between Fe depletion, either by Fe chelation therapy (14) or blood donation (15–17), and a decreased risk of CVD. Conversely, other cross-sectional (18, 19) and longitudinal (20) studies have reported a lack of an association. Most studies evaluating these relationships to date have been conducted in populations with a relatively high prevalence of chronic diseases. As a result, it remains uncertain if the observed variability in Fe status contributes to the onset of these diseases or is a consequence of the diseases themselves, particularly since many commonly used Fe status biomarkers (e.g., ferritin) can be elevated in response to inflammation.

Mendelian randomization (MR) is a statistical tool that uses genetic variation to explore causal effects of a risk factor on a health outcome in the presence of unknown confounders under three key assumptions: the genetic instruments selected as instrumental variables (IVs) must 1) reliably predict the exposure, 2) only be associated with the outcome through its association with the exposure, and 3) not be associated with confounders of the exposure-outcome association (**Figure S1**). To date, few MR studies evaluating the effects of Fe status on CVD risk have been published and these have found higher genetically predicted Fe status to be associated with a decreased risk of atherosclerotic disease (21–23) and hypercholesteremia (24) and an increased risk of ischemic stroke (IS) (25) and venous thromboembolism (VTE) (22). These MR studies used three genetic variants in Fe-regulatory genes as IVs selected from a European genome-wide association study (GWAS) of Fe biomarkers in < 50,000 individuals (26). However, the latest GWAS of Fe status biomarkers has reached a sample size of up to 257,953 and increased the number of genome-wide significant loci from 11 to 123 (27). Additionally, the IVs used in published studies do not satisfy all MR assumptions as these have been reported to have genome-wide significant associations with known confounders, including cholesterol, blood pressure, and body mass index (BMI). Consequently, it is unknown if the reported associations are confounded by pleiotropic effects of the IVs on CVD risk factors.

In depth analyses using the recently discovered genome-wide significant loci of Fe status as IVs are needed to obtain reliable effect estimates of Fe status on CVD risk. Our aim was to investigate the associations between genetically predicted Fe status and risk of ischemic stroke (IS) and coronary heart disease (CHD) by employing an MR framework that controls for CVD risk factors. A secondary aim was to investigate the pathways by which Fe status influences CVD risk utilizing mediation analyses.

## Methods

The work presented was performed using publicly available summary-level data from published GWAS. All analyses were conducted using R version 4.2.2 (The R Foundation for Statistical Computing). Results are reported per STROBE-MR recommendations (**Table S1**).

### Exposure data source

Summary-level data were obtained from the largest Fe GWAS available to date, which consisted of a meta-analysis of GWAS of six European populations (DeCODE, INTERNAL, SardiNIA, Danish Blood Donor Study (DBDS), Trøndelag Health Study (HUNT), and Michigan Genomics Initiative (MGI), for four Fe status biomarkers (serum Fe (SI), transferrin saturation (TSAT), serum ferritin (SF), and total Fe binding capacity (TIBC)) (27) (**Table 1**). Descriptive characteristics of the populations studied, the biomarker quantification methods utilized, and the data sources are outlined in **Table S2**. The physiological significance of the Fe status indicators is described in **Table S3**.

**Table 1.** Summary of exposure and outcome genome-wide association studies (GWAS) datasets.

|  | **Data Source** | **Source ID** | **Consortium** | **Variable type** | **Sample Size/**<br>**Cases/controls** | **Population** | **Sex** | **PMID** | **Year** |
| --- | --- | --- | --- | --- | --- | --- | --- | --- | --- |
| **Iron status** |  |  |  |  |  |  |  |  |  |
| TSAT | NTNU Open Research Data | 10.18710/S9TJEL | HUNT, MGI, deCODE, INTERVAL | Continuous | 198,516 | EUR | M/F | 35710628 | 2022 |
| SI | NTNU Open Research Data | 10.18710/S9TJEL | HUNT, MGI, deCODE, INTERVAL, SardinIA | Continuous | 236,612 | EUR | M/F | 35710628 | 2022 |
| TIBC | NTNU Open Research Data | 0.18710/S9TJEL | HUNT, MGI, deCODE, INTERVAL, SardinIA | Continuous | 208,422 | EUR | M/F | 35710628 | 2022 |
| Ferritin | NTNU Open Research Data o | 10.18710/S9TJEL | HUNT, MGI, deCODE, INTERVAL, DBDS | Continuous | 257,953 | EUR | M/F | 35710628 | 2022 |
| **Cardiovascular Disease** |  |  |  |  |  |  |  |  |  |
| CHD | IEU OpenGWAS | ieu-a-7 | CARDIoGRAM-plusCD4 | Binary | 60,801 / 123,504 | Mixed (76% EUR) | M/F | 26343387 | 2015 |
| CES | IEU OpenGWAS | ebi-a-GCST006910 | MEGASTROKE | Binary | 7,193 / 406,111 | EUR | M/F | 29531354 | 2018 |
| LAS | IEU OpenGWAS | ebi-a-GCST006907 | MEGASTROKE | Binary | 4,373 / 406,111 | EUR | M/F | 29531354 | 2018 |
| SVS | IEU OpenGWAS | ebi-a-GCST006909 | MEGASTROKE | Binary | 5,386 / 406,111 | EUR | M/F | 29531354 | 2018 |
| IS | IEU OpenGWAS | ebi-a-GCST006908 | MEGASTROKE | Binary | 34,217 / 406,111 | EUR | M/F | 29531354 | 2018 |
| AF | IEU OpenGWAS | ebi-a-GCST006061 | AFGen, BroadAF, and UKBB | Binary | 55,114 / 482,295 | EUR | M/F | 29892015 | 2018 |
| **Blood lipids** |  |  |  |  |  |  |  |  |  |
| TG | Neale Lab (R2) | 30870_irnt | UKBB | Continuous | 343,992 | EUR | M/F | NA | 2018 |
| TC | Neale Lab (R2) | 30690_irnt | UKBB | Continuous | 344,278 | EUR | M/F | NA | 2018 |
| LDL | Neale Lab (R2) | 30780_irnt | UKBB | Continuous | 343,621 | EUR | M/F | NA | 2018 |
| HDL | Neale Lab (R2) | 30760_irnt | UKBB | Continuous | 315,133 | EUR | M/F | NA | 2018 |
| Apo B | Neale Lab (R2) | 30640_irnt | UKBB | Continuous | 342,590 | EUR | M/F | NA | 2018 |
| Apo A | Neale Lab (R2) | 30630_irnt | UKBB | Continuous | 313,387 | EUR | M/F | NA | 2018 |
| **Blood Pressure** |  |  |  |  |  |  |  |  |  |
| DBP | Neale Lab (R2) | 4079_irnt | UKBB | Continuous | 340,162 | EUR | M/F | NA | NA |
| SBP | Neale Lab (R2) | 4080_irnt | UKBB | Continuous | 340,159 | EUR | M/F | NA | NA |

| **Anthropometric Measure** |  |  |  |  |  |  |  |  |  |
| --- | --- | --- | --- | --- | --- | --- | --- | --- | --- |
| BMI | Neale Lab (R2) | 21001_irnt | UKBB | Continuous | 359,983 | EUR | M/F | NA | NA |
| **Inflammatory Marker** |  |  |  |  |  |  |  |  |  |
| IL-6 | IEU OpenGWAS | ebi-a-GCST90012005 | 11 EUR cohorts | Continuous | 21,758 | EUR | M/F | 33067605 | 2020 |

AF, atrial fibrillation; Apo-A, apolipoprotein-A; Apo-B, apolipoprotein-B; BMI, body mass index; CARDIoGRAMplusC4D, Coronary ARtery DIsease Genome wide Replication and Meta-analysis plus The Coronary Artery Disease Genetics; CES, cardioembolic ischemic stroke; CHD, coronary heart disease; DBDS, Danish Blood Donor Study; DBP, diastolic blood pressure; EUR, European; Fe, iron; HDL, high density lipoprotein cholesterol; HUNT, Trøndelag Health Study; IL-6, interleukin-6; IS, ischemic stroke (all-types); LAS, large artery atherosclerotic ischemic stroke; LDL, low density lipoprotein cholesterol; M/F, males/females; MGI, Michigan Genomics Initiative; SBP, systolic blood pressure; SVS, small vessel ischemic stroke; TC, total cholesterol; TG, triglycerides; TIBC, total iron binding capacity; TSAT, transferrin saturation; UKBB, UK Biobank

### Outcome data sources

Detailed information on the outcome data sources used in this study are shown in **Table 1** and **Table S4**.

#### Cardiovascular disease risk factors

Known CVD risk factors were selected to 1) investigate the causal effect of Fe on the risk factor using two-sample univariate MR and 2) to investigate the confounding and/or mediating effects of the significant risk factors on the associations of genetically predicted Fe status and CVD using multivariate MR (MVMR). The risk factors selected were blood lipids (high density lipoprotein cholesterol (HDL), low density lipoprotein cholesterol (LDL), total cholesterol (TC), triglycerides (TG), apolipoprotein A (Apo-A), apolipoprotein B (Apo-B), blood pressure (diastolic blood pressure (DBP) and systolic blood pressure (SBP)), body mass index (BMI), and an inflammatory marker (interleukin-6 (IL-6)). Summary statistics for blood lipids, blood pressure outcomes and BMI were obtained from the UK Biobank and GWAS data for IL-6 were from a meta-analysis of eleven independent European cohorts (28).

#### Cardiovascular diseases

CHD, all-cause IS, and IS subtypes were selected as primary CVD outcomes. CHD summary statistics data were obtained from the CARDIoGRAMplusC4D (Coronary ARtery DIsease Genome wide Replication and Meta-analysis (CARDIoGRAM) plus The Coronary Artery Disease (C4D) Genetics) 1000-Genomes GWAS meta-analysis (29), which consisted of a GWAS meta-analysis of CHD in forty-eight multi-ethnic populations. Of the total sample size, 76% of the participants included were of European descent. Summary-level data for IS and IS subtypes (cardioembolic (CES), large artery atherosclerosis (LAS), small-vessel (SVS)) were obtained from a GWAS meta-analysis in seventeen European populations led by the MEGASTROKE consortium (30). We were unable to rule out potential sample overlap between the MEGASTROKE and the exposure data because both GWAS meta-analyses involved individuals from deCODE. To mitigate the risk of type I error (31), we tested an additional outcome closely related to IS, atrial fibrillation (AF), to validate our findings. Summary-level GWAS data for AF were obtained from a GWAS meta-analysis of European consortiums including Atrial Fibrillation Genetics consortium (AFGen) and the Broad AF study (32).

### Instrumental variable selection

Independent SNPs (*r*^2^ < 0.001) associated with each Fe trait at genome-wide significance level (*P* < 5×10^−8^) were extracted from the Fe GWAS meta-analysis (27). Rare SNPs with a minor allele frequency (MAF) < 1% in Europeans were excluded. SNPs defined as being ambiguous with intermediate allele frequencies were removed. Additionally, to further minimize the potential for pleiotropy, SNPs that had direction of effects that were not consistent with systemic Fe status were removed (i.e., higher Fe status results in increased TSAT, serum Fe and ferritin, and decreased TIBC). Lastly, SNPs that were not genotyped in the outcome dataset were replaced by proxy SNPs if available or removed if no proxy SNP was found. Proxy SNPs were defined as a SNP in linkage disequilibrium (LD; *r*^2^ > 0.8) with the genetic instrument in a European reference population (**Table S5**).

The strength and validity of the genetic instruments were evaluated by calculating the variance in the Fe trait explained by a SNP (*R*^2^) and the *F*-statistic. The *R*^2^ for each SNP was calculated using the equation *R*^2^ = 2β^2^ MAF (1-MAF), where β is the regression coefficient from the SNP-Fe trait association from the GWAS, and MAF is the minor allele frequency for that SNP. The *F*-statistic was calculated using the equation *F*-statistic = (*n*-*k*-1/*k*)(*R*^2^ _instrument_ / 1-*R*^2^ _instrument_), where *n* is the population sample size, *k* is the number of SNPs in the instrument, and *R*^2^ _instrument_ is the sum of the *R*^2^ for each SNP included in the instrument (33). We considered the MR standard *F*-statistic > 10 to indicate adequate instrument strength (33). The SNPs selected as instrumental variables and their respective *R*^2^ and *F*-statistic are shown in **Table S5**. The *R*^2^ _instrument_ and the *F*-statistic of the instruments were calculated after data harmonization for all four Fe biomarkers for each outcome evaluated as shown in **Table S6**.

### Mendelian randomization

We developed a workflow combining variations of the MR method (**Figure 1**). Initial analyses consisted of univariate MR analyses to identify major CVD risk factors that are influenced by Fe traits (**Figure 1**; step 1a) and to evaluate the total effect of Fe traits on CVD outcomes (**Figure 1**; step 1b). Variables meeting the selection criteria were carried over to step 2, which consisted of MVMR analyses to determine the direct effects of Fe traits on CVD outcomes conditioning on major CVD risk factors (**Figure 1**; step 2). Lastly, CVD risk factors satisfying the selection criteria were carried over to step 3, where these were evaluated as mediators of the Fe trait-CVD associations and the indirect effects of Fe traits on CVD were calculated (**Figure 1**; step 3). The detailed MR models implemented, and the variables tested at each step are presented in **Figure S2**.

**Figure 1.**
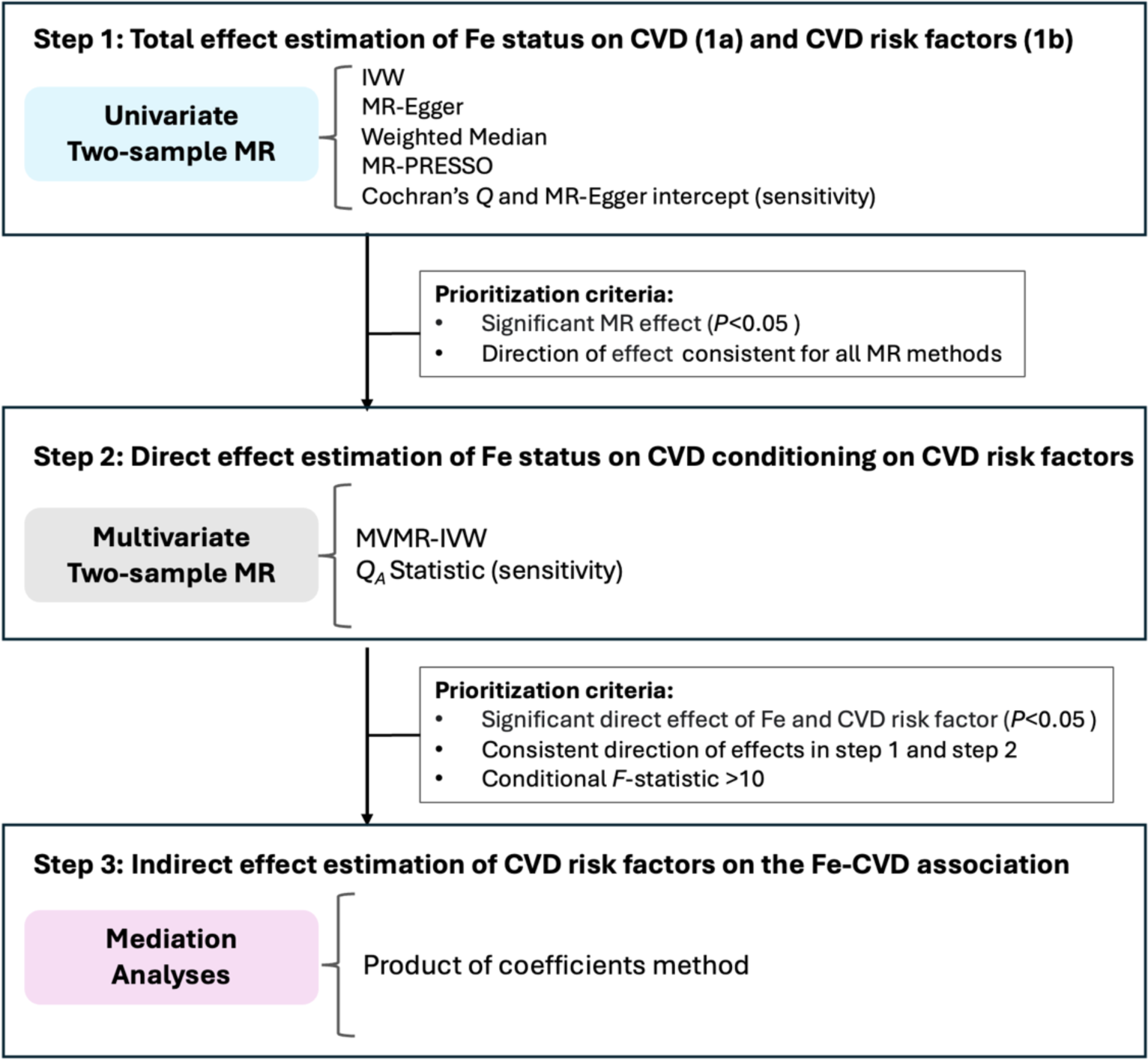
Study design and workflow. CVD, cardiovascular disease; Fe, iron; IVW, inverse variance weighted; MR, Mendelian Randomization; MR-PRESSO, Mendelian randomization pleiotropy residual sum and outlier test; MVMR, multivariate mendelian randomization.

#### Univariate Mendelian randomization

We performed three main MR estimations for each exposure-outcome test, which included the inverse variance weighted (IVW) method under multiplicative random effects (34), the MR-Egger method, and the weighted median (WM) method. The IVW method requires either all SNPs used as IVs to be valid instruments or that there is balanced horizontal pleiotropy, whereas the MR-Egger and weighted median are more robust to bias introduced by weak IVs; the MR-Egger is less susceptible to bias introduced by horizontal pleiotropy (35) and the weighted median (WM) provides an unbiased estimate if <50% of the SNPs used as IVs present evidence of pleiotropy (36). When significant evidence of pleiotropy was detected (described below under “sensitivity analyses”), the WM and MR-Egger estimates were prioritized for interpretation of results. Univariate MR analyses were performed using the *‘TwoSampleMR’* R package (37). MR estimates are presented as the β and standard error (SE) per 1-SD unit change in the Fe status biomarker for continuous outcomes, and as odds ratio (OR) and 95% CI per 1-SD unit change in the Fe status biomarker for dichotomous outcomes.

#### Multivariate Mendelian randomization

MVMR is an extension of the two-sample univariate MR method that can be employed to estimate the effect of an exposure on an outcome while controlling for confounders (38). We performed MVMR on CVD outcomes meeting our prioritization criteria conditioning on one CVD risk factor at a time. MVMR estimates were calculated using the IVW method and all MVMR analyses were performed using the ‘*MVMR*’ R packages (39, 40), which extend the typical MR main and sensitivity tests (IVW and Cochran’s *Q*) to MVMR analyses. The conditional *F*-statistic, which accounts for the association between each SNP with other exposures included in the estimation, was calculated to assess instrument strength.

#### Mediation analysis

Mediation analyses were conducted to assess the mechanisms by which Fe status influences CVD outcomes. Effect estimates obtained from the univariate MR (total effects of Fe on CVD (**Figure 1**; step1a) and CVD risk factor (**Figure 1**; step1b)) and MVMR (direct effects of Fe on CVD conditioning on a CVD risk factor (**Figure 1**; step 2)) were used to calculate the indirect effects of Fe status on a CVD outcome using the product of coefficients method (41). This method was selected as the outcomes evaluated are considered rare with a prevalence <10% (42). The SE and 95% CI were calculated using the Delta method (43). The percent of effect mediated by a CVD risk factor was calculated as the estimated indirect effect divided by the total effect from univariate MR x 100 (43). Mediation estimates are presented as the log odds ratio, SE and 95% CI.

### Sensitivity Analyses

Pleiotropy refers to a phenomenon in which a variant has causal effects on more than one phenotype. In the context of MR, significant bias is introduced when there is evidence of pleiotropy within the genetic instruments (i.e., if the MR assumption 3 is not met). We evaluated the presence of directional pleiotropy using the Cochran’s *Q* statistic and the MR-Egger intercept for the univariate MR analyses. Additionally, we performed the MR pleiotropy residual sum and outlier (MR-PRESSO) test using the *‘MRPRESSO’* package in R, which identifies pleiotropic outliers (44). When outliers were detected, the MR-PRESSO outlier-corrected effect estimates were calculated and presented. For MVMR analyses, heterogeneity was evaluated with the *Q_A_* statistic. Lastly, we evaluated potential pleiotropic effects of the SNPs used as genetic instruments using the online tools LDtrait (45) and PhenoScanner (46, 47). We searched for Fe-associated SNPs or SNPs in LD (*r*^2^ > 0.8) with these SNPs that had genome-wide significant associations (*P* < 5×10^−8^) with CVD risk factors (such as blood lipids, BMI, and blood pressure) or with CVD outcomes (such as CHD, IS, and AF) (**Table S5**). SNPs previously reported to have a genome-wide significant association with a CVD outcome were excluded in sensitivity analyses for MVMR.

### Power Calculations

Statistical power for binary outcomes (i.e., CVD outcomes) was calculated using the online tool for power calculations for MR (48) and for continuous outcomes (i.e., CVD risk factors) using published methods and R code (49). Based on our strongest instrumental variable with an *R*^2^ of 0.055 at an α=0.05, our analyses were sufficiently powered (>80%) to detect an odds ratio ≤ 0.94 or ≥ 1.06 for CHD, ≤ 0.93 or ≥ 1.07 for IS, ≤ 0.81 or ≥ 1.19 for CES, ≤ 0.83 or ≥ 1.17 for LAS, ≤ 0.85 or ≥ 1.15 for SVS, and ≤ 0.94 or ≥ 1.06 for AF using two-sample MR. Detailed power calculations for each Fe exposure-outcome combination are shown in **Table S7**.

## Results

### Univariate Mendelian randomization

We used common independent SNPs (**Table S5**) explaining 5.5%, 3.9%, 3.2% and 1.5% of the variance in TSAT, TIBC, serum Fe and ferritin, respectively. Instrument *F*-statistic for all four Fe biomarkers ranged from 109 to 1024 after harmonization of exposure and outcome data (**Table S6**).

The MR estimates for the effects of genetically predicted Fe status on CVD risk factors are shown in **Tables S8-S11**. For the Fe-blood lipids, Fe-BP, and Fe-BMI analyses the WM and MR-Egger estimates were prioritized for interpretation of results as the Cochran’s *Q* statistic demonstrated evidence of pleiotropy between genetic instruments (*P* < 0.05); little to no evidence of pleiotropy was evident from the MR-Egger intercept test (*P* > 0.05). For the Fe-IL-6 analyses, no evidence of heterogeneity was observed (*P* > 0.05 for the Cochran’s *Q* statistic and MR-Egger intercept); thus, the IVW MR estimates were taken into consideration.

Higher genetically predicted TSAT, indicative of higher Fe status, was associated with lower LDL cholesterol (WM: β = −0.11; SE = 0.01, *P* = 3.9×10^−14^), TC (WM: β = −0.10; SE = 0.02, *P* = 5.3×10^−10^), and Apo-B (WM: β = −0.10; SE = 0.01, *P* = 9.2×10^−13^), and with higher Apo-A (WM: β = 0.02; SE = 0.009, *P* = 0.007), and triglycerides (WM: β = 0.03; SE = 0.009, *P* = 0.002). No evidence of an effect was observed for HDL (WM: *P* = 0.5), BMI (WM: *P* = 0.1), or IL-6 (IVW: *P* = 0.2). With respect to blood pressure outcomes evaluated, higher genetically predicted TSAT was associated with higher DBP (WM: β = 0.06; SE = 0.01, *P* = 1.22×10^−9^), but was not associated with SBP (WM: *P* = 0.7). Consistent effects on all CVD risk factors except for BMI were observed when serum Fe and TIBC were evaluated as exposures (**Tables S8-S10**). While genetically predicted TSAT and serum Fe were not associated with BMI, higher TIBC (indicative of lower Fe status) was associated with a higher BMI (WM: β = 0.03; SE = 0.009, *P* = 0.01). Overall, the direction of effects for ferritin as the exposure were consistent with the other Fe biomarkers, but weaker associations were observed due to the lower statistical power of its IVs (**Table S11**).

The MR-PRESSO analyses identified possible pleiotropic outliers for all CVD risk factors evaluated except for IL-6 (**Tables S8-S11)**. For analyses of TIBC as the exposure, no change was observed with respect to significance and direction of effect after implementation of the outlier correction (**Table S10**), but few differences were observed for analyses of TSAT and serum Fe as the exposures (**Tables S8 and S9**). The MR-PRESSO outlier-corrected estimates suggested a significant association of lower genetically predicted TSAT (WM: β = −0.03; SE = 0.009, *P* = 0.01) and serum Fe (WM: β = −0.04; SE = 0.01, *P* = 0.01) with a higher BMI, but a null effect on the blood lipids and blood pressure (**Tables S8 and S9**). Caution should be taken when interpreting these estimates as the lack of an effect may have resulted from a weak instrument and a lower statistical power after the removal of SNPs.

The MR OR per 1-SD unit increase in TSAT, serum Fe, TIBC and ferritin are presented in **Table 2**. Univariate MR analyses showed evidence for a causal effect of higher Fe status on CVD outcomes, but the direction of effect differed by outcome. Higher genetically predicted Fe status, as evidenced by a higher TSAT, serum Fe, or lower TIBC, was associated with a greater risk of CES, a lower risk of CHD, and was not associated with all-cause IS or SVS. Lower genetically predicted TIBC, indicative of higher Fe status, was associated with a lower risk of LAS. Using MR-PRESSO, outliers were detected for the associations of TSAT and TIBC with CHD. Nonetheless, outlier-corrected estimates and *P*-values were in agreement with the other MR methods (**Table 2**). Analyses for AF appeared to have significant heterogeneity (*P*-value for Cochran’s *Q* statistic < 0.05); thus, WM and MR-PRESSO were prioritized for interpretation. Consistent with the detrimental effects of higher Fe status on CES, we found greater genetically predicted Fe status, as evidenced by higher TSAT and serum Fe and lower TIBC, to be associated with an increased risk of AF (**Table 2**).

**Table 2.**
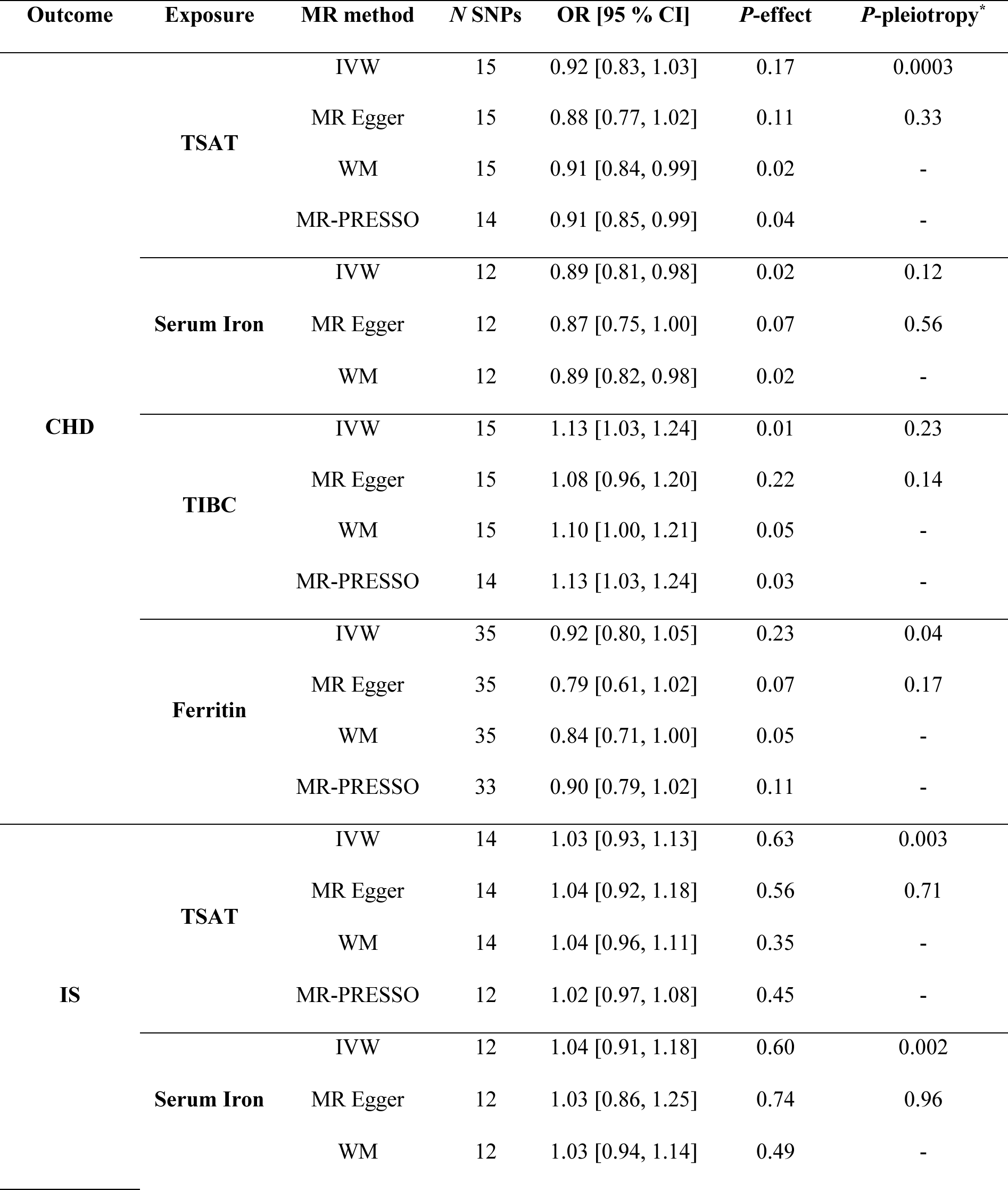
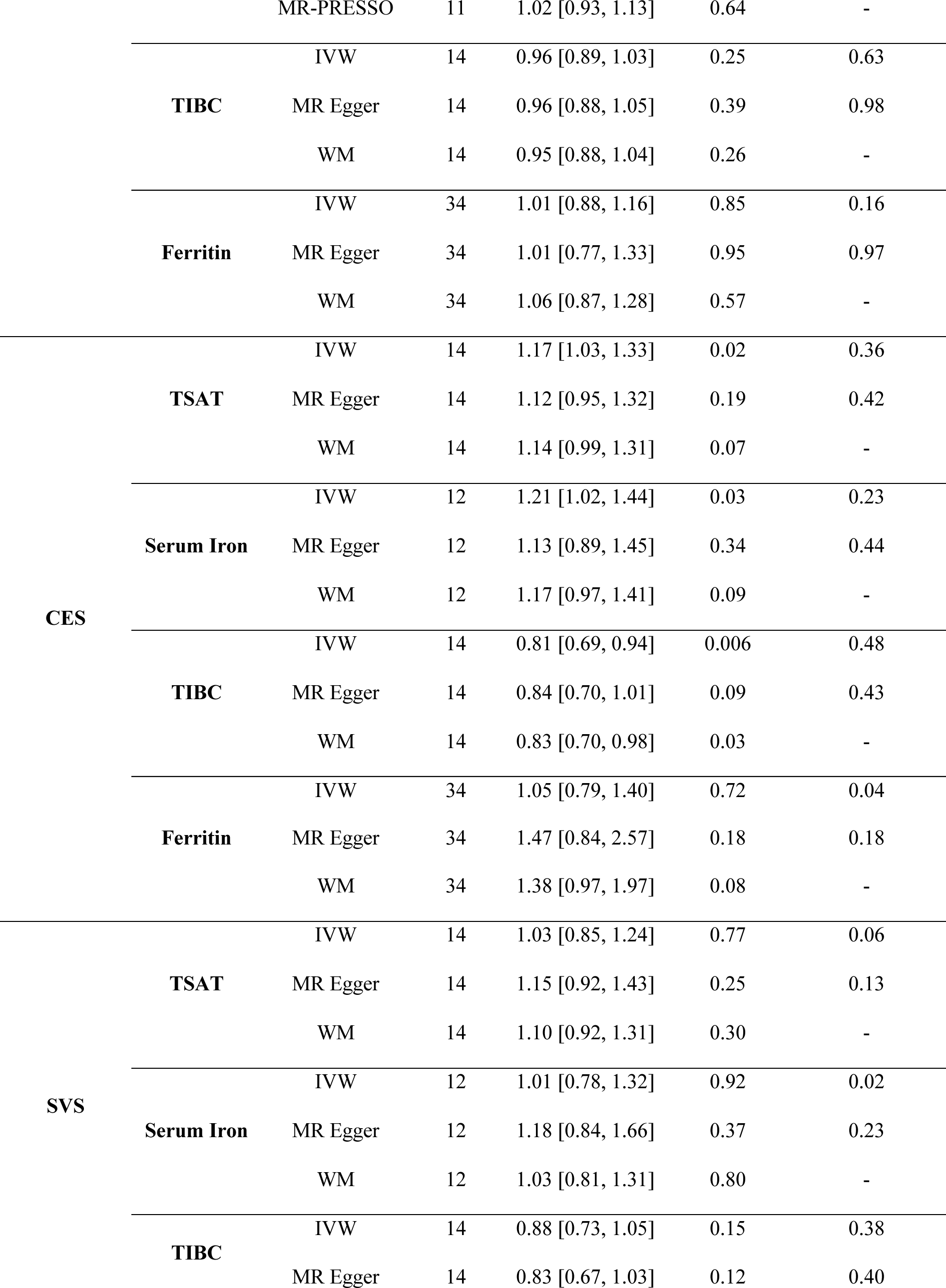
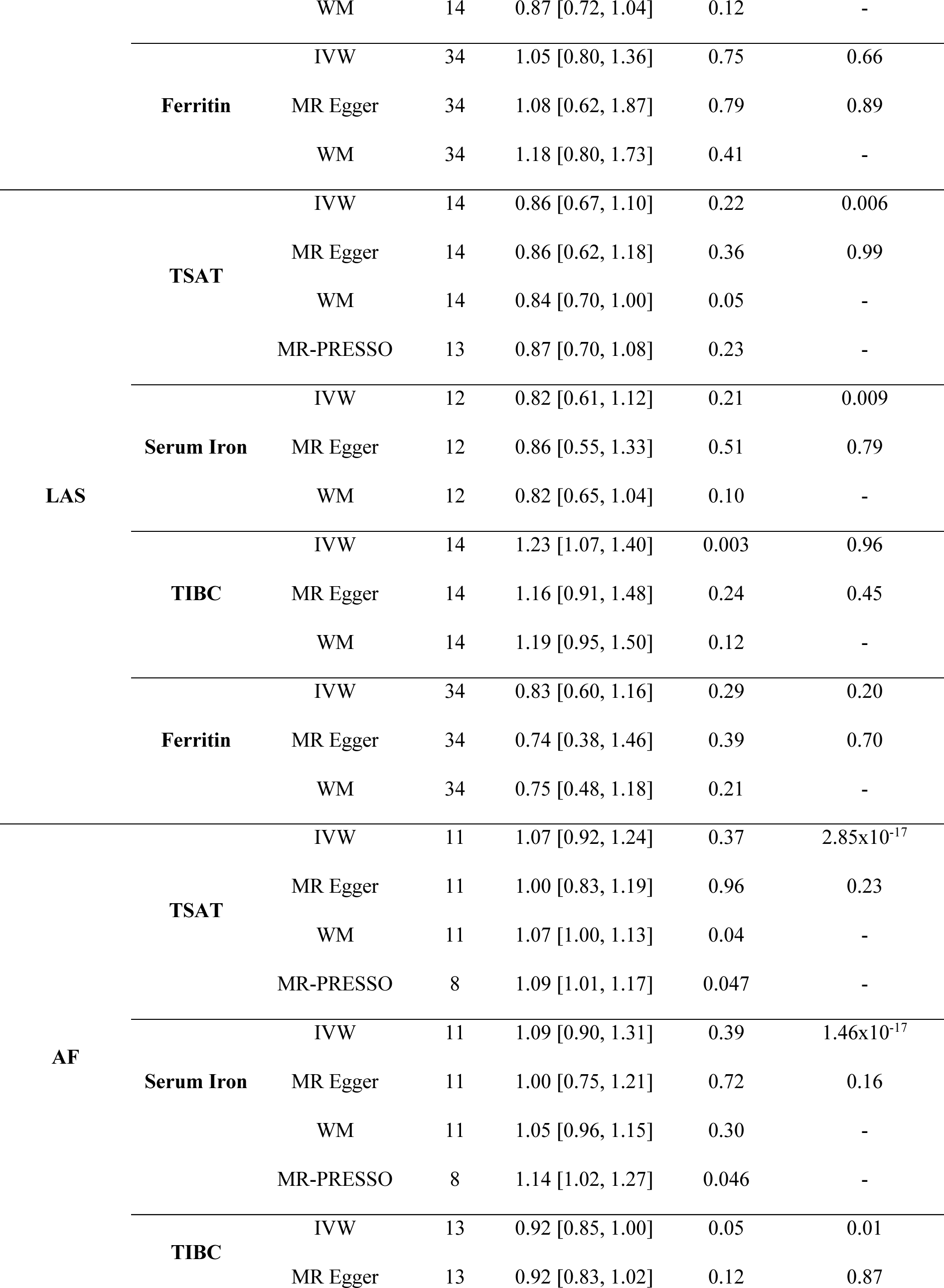
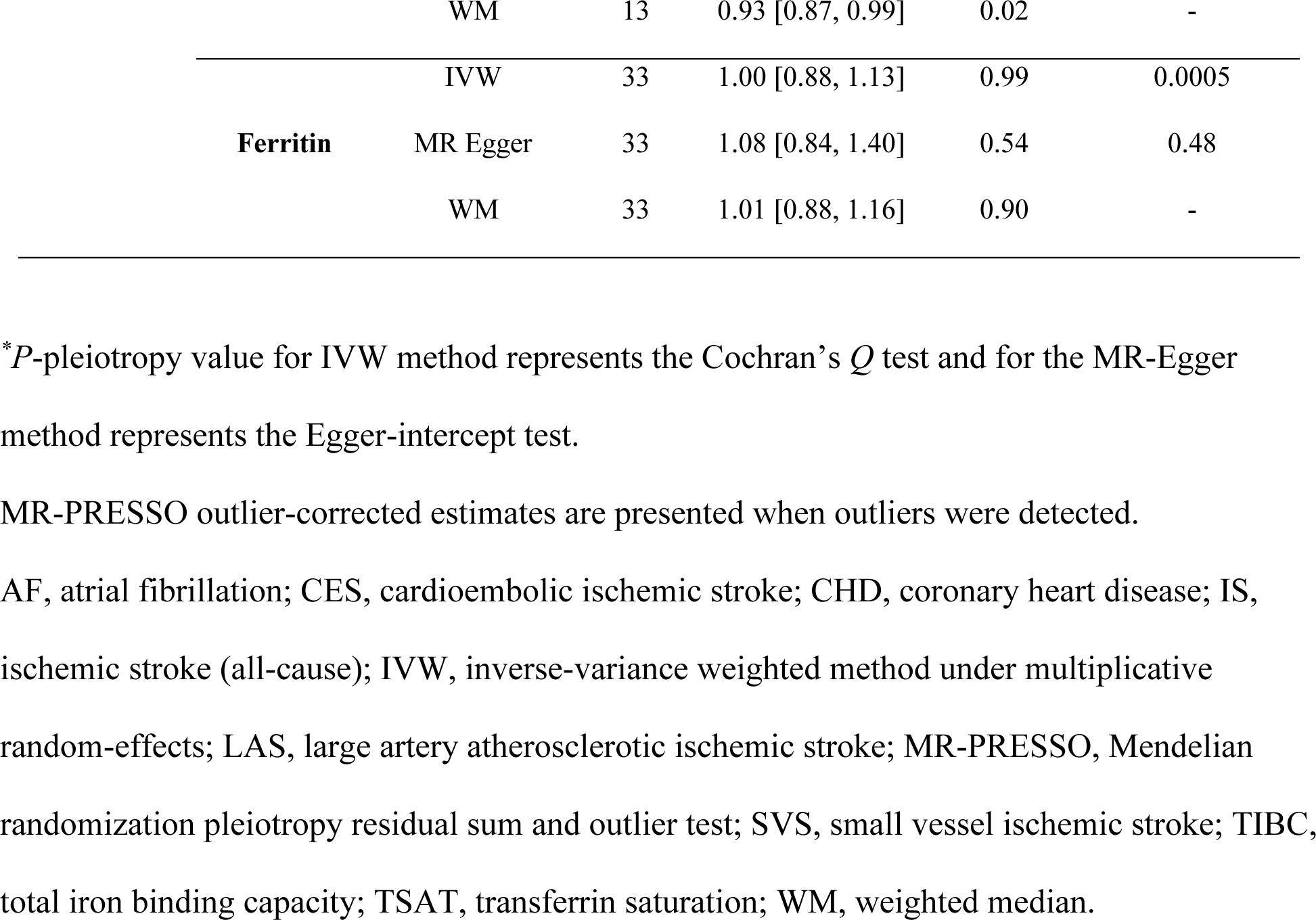
Univariate two-sample Mendelian randomization analyses evaluating the associations of genetically predicted iron status and cardiovascular disease outcomes.

### Multivariate Mendelian randomization

We carried out MVMR utilizing selected variables meeting the prioritization criteria from the univariate MR analyses. Because the power of the genetic instrument is greatly reduced when conditioning on additional exposures using MVMR, we excluded ferritin from the MVMR analyses given that the genetic instruments for ferritin only explain a small proportion of its variance. For all MVMR models of TSAT, serum Fe or TIBC as exposures, the conditional *F*-statistic was > 10, demonstrating that weak instrumental bias is unlikely to be present (**Tables S12-S14**).

MVMR analyses provided evidence for a causal effect of higher Fe status on CES after conditioning on CVD risk factors, and this observation was consistent across TSAT, serum Fe and TIBC as shown in **Figure 2**. The protective effect of higher Fe status on CHD and LAS observed in the univariate MR analyses was entirely nullified after controlling for blood lipids (**Figure 3**). The confounding effects of BMI on the analyses were not consistent throughout the three Fe status exposures. The protective effect of lower TIBC and higher TSAT on CHD remained significant when conditioning for BMI. However, the presumed effects serum Fe on CHD disappeared when conditioning on BMI (**Figure 3**).

**Figure 2.**
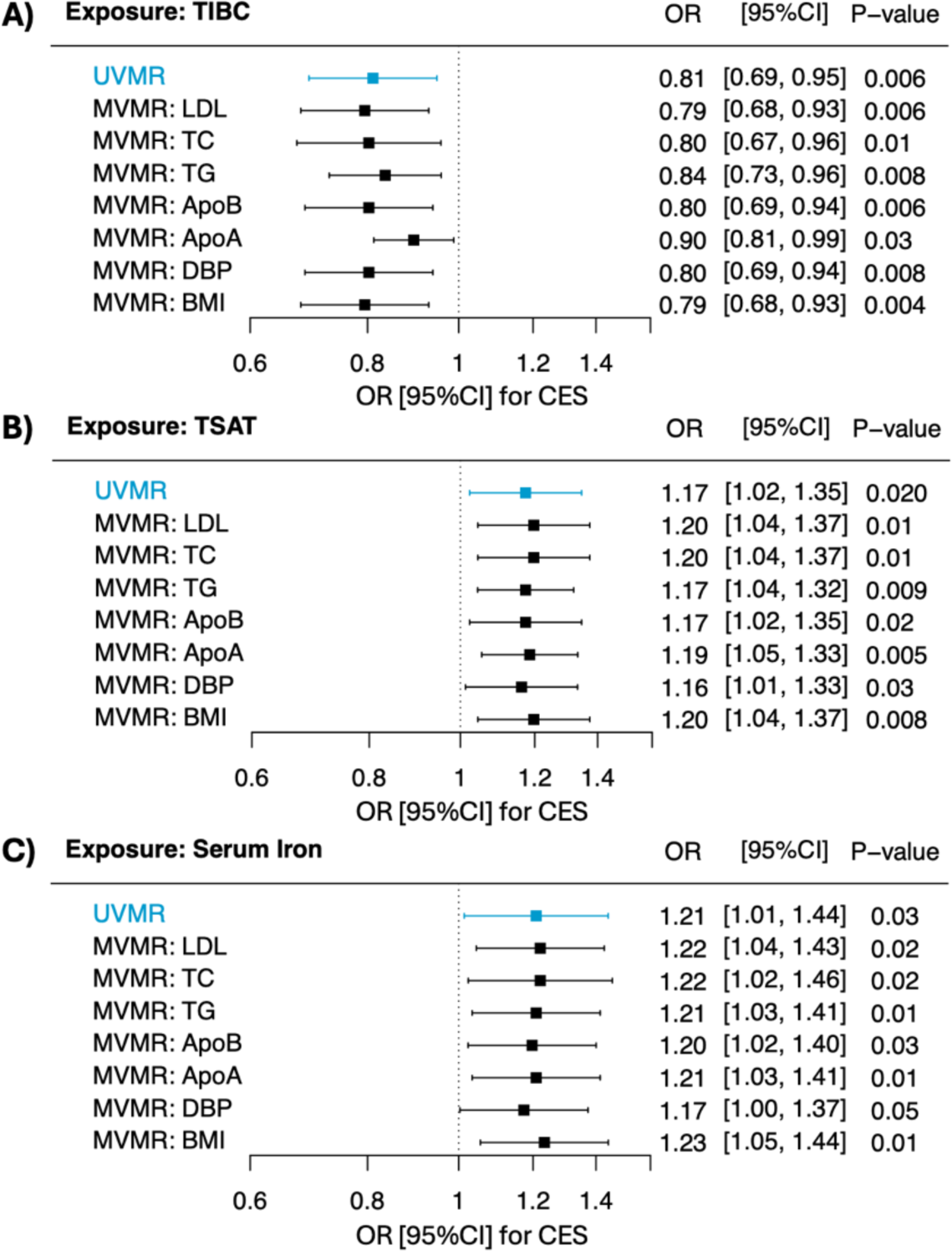
Associations between genetically predicted iron status and cardioembolic ischemic stroke using univariate and multivariate Mendelian randomization. MR estimates presented as odds ratio (OR) and 95% confidence intervals (CI) per 1-SD unit increase in the Fe exposure. Apo-A, apolipoprotein-A; Apo-B, apolipoprotein-B; BMI, body mass index; CES, cardioembolic ischemic stroke; DBP, diastolic blood pressure; LDL, low density lipoprotein cholesterol; MVMR, multivariate Mendelian randomization; TC, total cholesterol; TG, triglycerides; UVMR, univariate Mendelian randomization.

**Figure 3.**
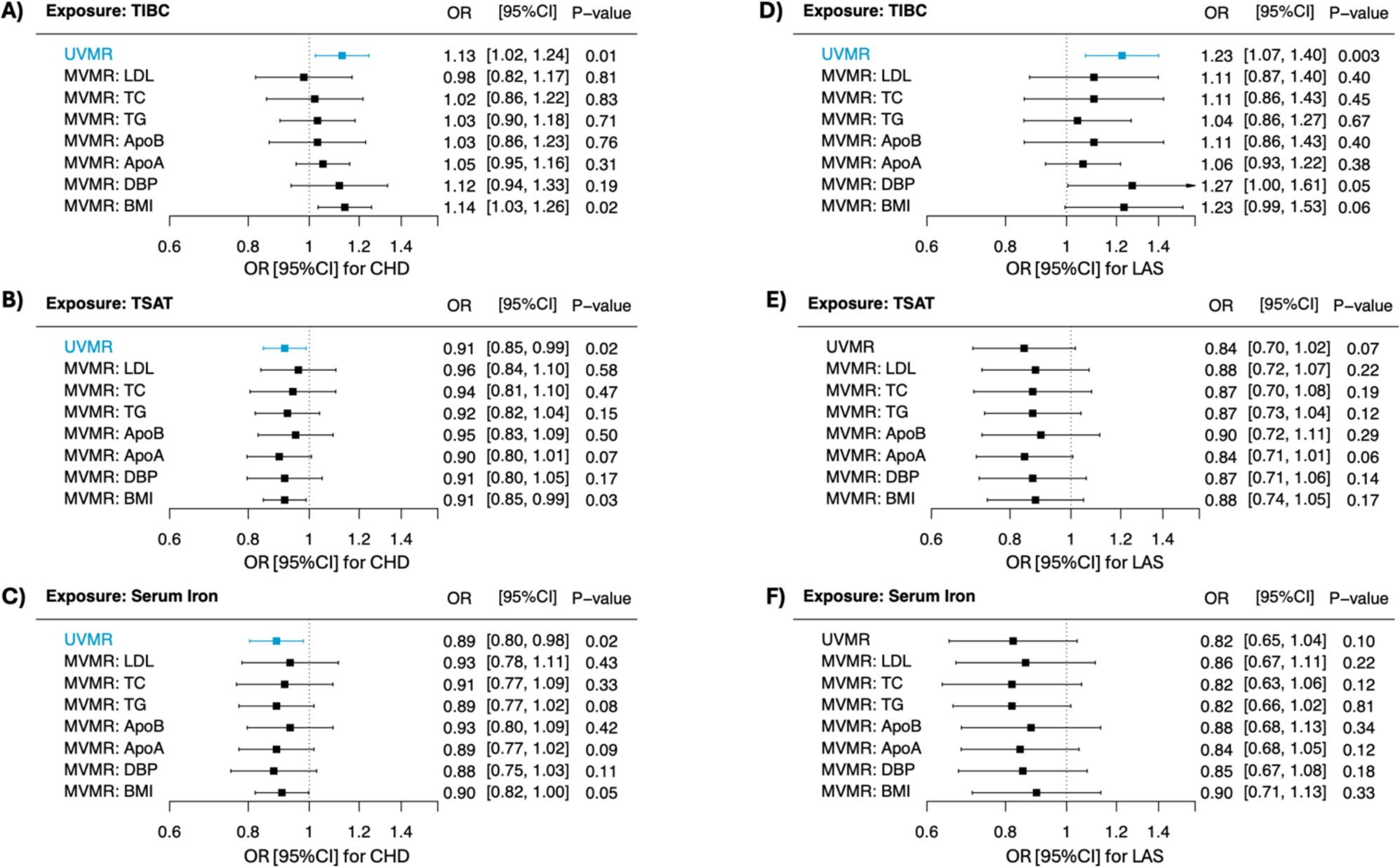
Associations between genetically predicted iron status and atherosclerotic cardiovascular disease outcomes using univariate and multivariate Mendelian randomization. MR estimates presented as odds ratio (OR) and 95% confidence intervals (CI) per 1-SD unit increase in the Fe exposure. Apo-A, apolipoprotein-A; Apo-B, apolipoprotein-B; BMI, body mass index; DBP, diastolic blood pressure; CHD, coronary heart disease; LAS, large artery atherosclerotic ischemic stroke; LDL, low density lipoprotein cholesterol; MVMR, multivariate Mendelian randomization; TC, total cholesterol; TG, triglycerides; UVMR, univariate Mendelian randomization.

Evaluation of heterogeneity using the *Q_A_* statistic showed little to no heterogeneity in the analyses evaluating CES as the outcome (*P*-heterogeneity > 0.05) (**Table S12**). Substantial heterogeneity was noted in the MVMR analyses for CHD and LAS (*P*-heterogeneity < 0.05) (**Tables S13 and S14**). From our LDlink and PhenoScanner searches we identified two SNPs that have been shown to be associated at genome-wide significance level with coronary artery disease and one SNP with venous thromboembolism (**Table S5**). Additional sensitivity analyses excluding these three SNPs did not affect the results, thus, demonstrating robustness of findings (**Table S15**).

### Mediation analyses

We conducted mediation analyses to evaluate the indirect effect of TSAT, serum Fe and TIBC on CES via selected mediators (DBP, BMI, LDL, TC, TG, Apo-B, Apo-A) using effect estimates derived from univariate MR and MVMR analyses. Significant mediation of the Fe status-CES relationship via DBP was observed consistently across the three Fe status exposures (**Table 3**). The estimated percent of the total effect of Fe status on CES mediated by DBP was 7.1, 8.0 and 8.8 % for serum Fe, TIBC and TSAT, respectively. The magnitude of mediation observed for BMI and blood lipid fractions evaluated was substantially lower (0-2 %) and was not statistically significant based on the wider 95% CI that crossed the null. Detailed calculations are shown in **Table S16**.

**Table 3.**
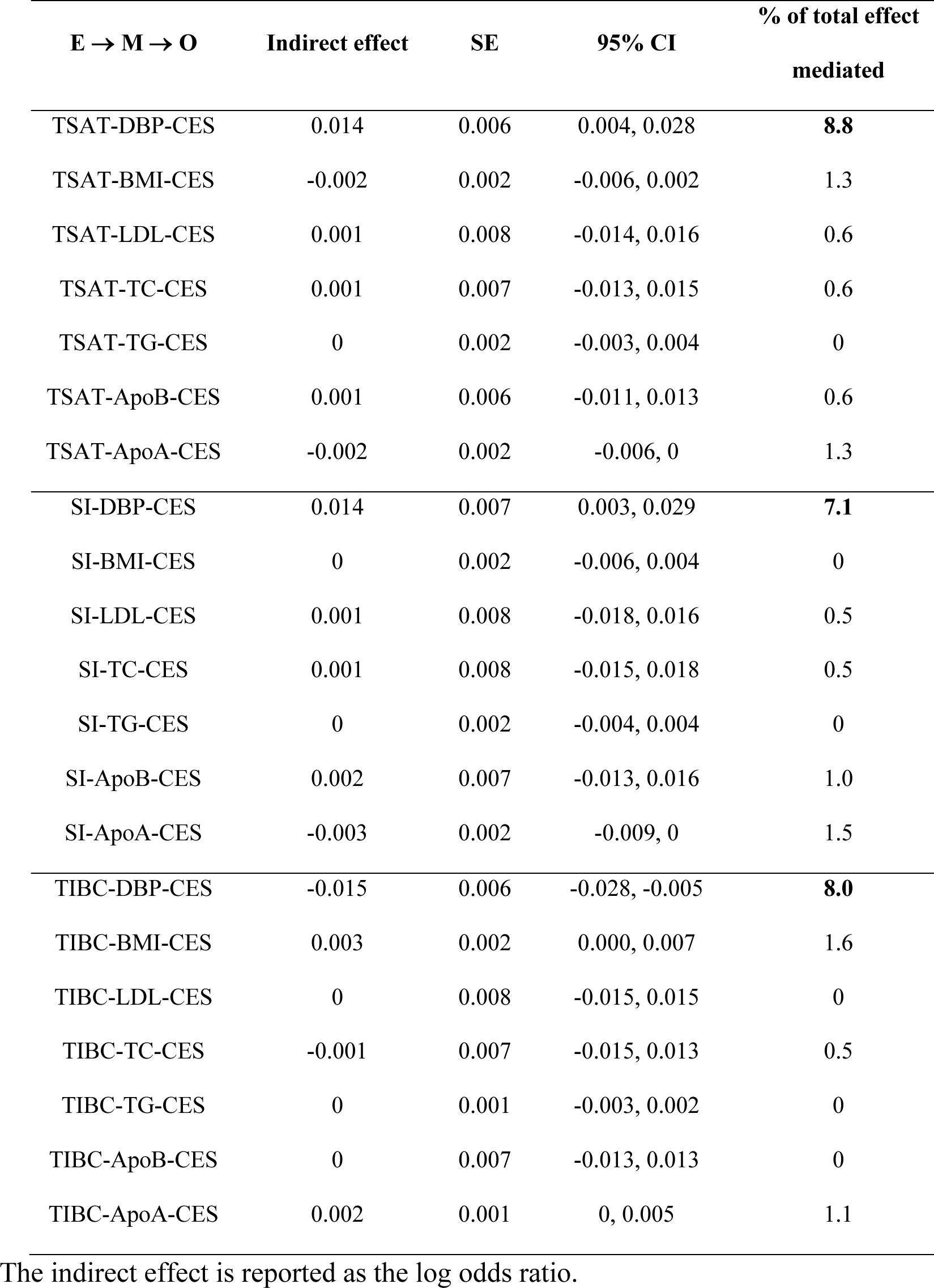
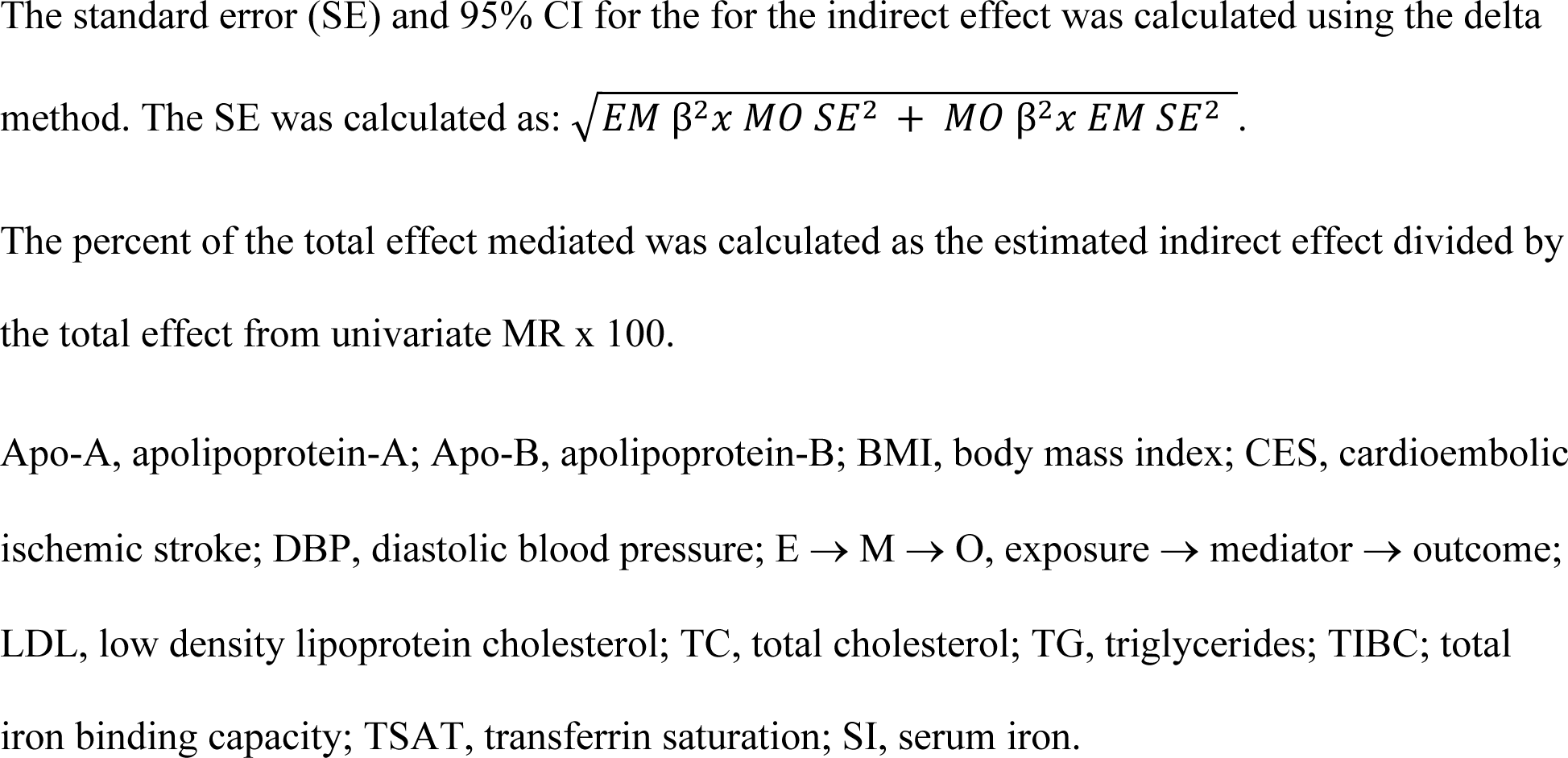
Indirect effects reported as log odds ratio of iron status on cardioembolic stroke via selected mediators.

## Discussion

To our knowledge, this is the first study to implement MVMR to evaluate the effects of Fe status on CVD outcomes while adjusting for potential confounding variables. Our findings revealed a noteworthy association between higher genetically predicted Fe status and an increased risk of CES that is independent of major CVD risk factors, including blood lipids, DBP, and BMI. Further employing mediation analyses, we found that DBP partially mediates the effect of Fe status on CES. Interestingly, while univariate MR initially suggested a protective effect of higher Fe status on CHD and LAS, our comprehensive MVMR analyses factoring CVD risk factors, revealed a complete annulment of this perceived protective effect.

Our analyses using univariate two-sample MR to evaluate the effects of Fe status on CVD outcomes were consistent with previous MR studies based on smaller-scale GWAS of Fe traits (21–25). First, we replicated the observed protective effect of higher Fe levels on atherosclerotic CVD outcomes, including CHD and LAS. We also replicated the analyses pertaining to CES indicating an adverse effect of higher Fe status on this outcome. Lastly, consistent with a previous MR study (50), we found an association between higher genetically predicted Fe status with an increased risk of AF. This finding holds particular relevance as AF is the most significant risk factor for CES (51), aligning consistently with the observed effects of Fe levels on CES.

We further evaluated the causal effects of Fe status on major CVD risk factors using univariate MR. We found, for the first time using causal inference methods, an association between higher genetically predicted Fe status and higher DBP. The lack on an association between Fe status and SBP may have resulted from a low statistical power as suggested by our power calculations. Few studies to date have evaluated the potential effects of Fe status on blood pressure. A phenome-wide association study implementing MR found higher Fe status to increase risk of hypertension at a nominal significance level (25). In agreement, observational studies have shown that higher serum ferritin is associated with an increased risk of hypertension in adult men (52, 53), and serum Fe was found to be positively associated with an increased risk of hypertension during pregnancy (54). We then evaluated the causal effects of Fe status on BMI and found lower Fe status, as evidenced by higher TIBC, to be associated with a higher BMI. This is consistent with observational studies reporting a relationship between obesity and Fe deficiency, which is believed to stem from an interplay between Fe regulation and adiposity (55). With respect to blood lipids, higher genetically predicted Fe status was associated with better lipid profiles as evidenced by lower LDL, TC, and Apo-B and higher Apo-A concentrations. While these results are in agreement with a previous MR study showing an association between genetically predicted higher serum Fe and lower LDL and TC levels, and a decreased risk of hypercholesteremia and hyperlipidemia (24, 56), we found evidence suggestive of substantial pleiotropy in the analyses. The observed heterogeneity likely results from the significant overlap between Fe and lipid IVs as evidenced by our sensitivity analyses showing a lack of an association after removal of pleiotropic outliers. Lastly, genetically predicted Fe status was not associated with IL-6 in our analyses. Nonetheless, future MR studies should evaluate this relationship in the opposite direction (i.e., the effects of IL-6 on Fe status) as it is well-established that inflammation, particularly via IL-6, is a negative regulator of Fe status (57).

Although the results from univariate MR analyses carry important implications, a major limitation is the potential bias introduced by pleiotropic effects of the genetic instruments as several Fe-associated SNPs are strongly associated with major CVD risk factors. To overcome this limitation, we performed MVMR analyses conditioning on selected CVD risk factors. Most notably, with these analyses we found that the effects of Fe status on CES were unaffected by adjustment for DBP, BMI, or blood lipids (LDL, TC, TG, Apo-B, Apo-A). This provides evidence in support of a causal effect of higher Fe status on CES incidence that is independent of major CVD risk factors. Available observational evidence evaluating the associations between Fe status and risk of stroke is inconclusive as reviewed (25). However, these contrasting observations may stem from differences between IS subtypes as evidenced by our opposing results when evaluating IS by subtype. This highlights the importance of evaluating IS with stratification by subtype.

We hypothesized that Fe’s detrimental effect on CES is at least partially mediated by CVD risk factors evaluated in this study. Using mediation analyses, we identified DBP as potential pathway through which elevated Fe levels contribute to an increased risk of CES. Accumulation of excess Fe, such that it increases the labile Fe pool, results in the production of highly reactive species, consequently inducing oxidative stress. It is known that oxidative stress contributes to the pathogenesis of hypertension (58), a common vascular risk factor for CES. Thus, Fe-induced oxidative stress may be a mechanism by which higher Fe levels influence CES development directly and indirectly via its effects on blood pressure. Blood lipids did not appear to play a mediating role in the relationship between Fe status and CES, suggesting that the effect of higher Fe status on CES risk is unlikely to operate via the blood lipid fractions evaluated.

In contrast to the robust evidence observed for CES, the apparent protective effect of higher Fe status on CHD and LAS was diminished after adjustment for CVD risk factors using MVMR, suggesting a lack of a direct effect of Fe status on CHD and LAS when accounting for major CVD risk factors. Of note, significant heterogeneity was observed in these analyses, particularly when evaluating CHD as the outcome. Thus, more investigations are needed to characterize the complex interplay between Fe, atherosclerosis, and subsequent CVD outcomes.

Our study has limitations that warrant attention. First, due to our reliance on summary-rather than individual-level data, we were unable to explore potential sex-based differences in the relationships between Fe status and CVD risk factors. This is particularly relevant to Fe and CVD as both Fe status markers and risk of CVD differ significantly between males and females. To our knowledge, there are no sex-stratified summary-level GWAS data from either Fe status or CVD that are publicly available and adequately powered to address these questions. However, it remains crucial to replicate these analyses in males and females separately. Furthermore, while we selected the best available outcome datasets that closely matched the population in the exposure dataset and that adjusted for similar covariates, the possibility of population stratification cannot be ruled out. Additionally, since our study was based in European populations, we are unable to extrapolate our results to non-European populations. Evaluating this question in other populations is of pressing need as risk of excess body Fe accumulation differs between populations, with individuals of East Asian ancestry presenting the greatest body Fe burden (59). Another important consideration is that MR methods estimate the lifetime effect of Fe status; thus, the magnitude of the MR estimates may be greater in our study than what would be observed clinically. This characteristic of MR holds implications for our study as Fe accumulation differs throughout life stages. For instance, risk of Fe overload in females escalates post-menopause due to the cessation of menses and the effect size of elevated Fe on disease development may differ accordingly. Lastly, all MR approaches implemented in this study assume linear associations. However, there is a possibility that the effect of Fe on some of the exposures evaluated is not linear, potentially masking some effects.

In conclusion, our findings underscore the significance of Fe status as a novel modifiable risk factor for CES. Additionally, when controlling for CVD risk factors, the presumed protective effect of higher Fe status on atherosclerotic heart disease outcomes was negated. Future studies are needed to evaluate possible sex and population differences in the effects of Fe status on CVD risk. Lastly, given the inherent limitations of MR methods, additional studies are needed to confirm then clinical utility of Fe status in predicting risk of CES.

## Abbreviations

AF: atrial fibrillation
Apo-A: apolipoprotein-A
Apo-B: apolipoprotein-B
BMI: body mass index
CARDIoGRAMplusC4D: Coronary ARtery DIsease Genome wide Replication and Meta-analysis plus The Coronary Artery Disease Genetics
CES: cardioembolic ischemic stroke
CHD: coronary heart disease
CVD: cardiovascular disease
DBDS: Danish Blood Donor Study
DBP: diastolic blood pressure;
Fe: iron
HDL: high density lipoprotein cholesterol
HUNT: Trøndelag Health Study
IL-6: interleukin-6
IS: all-cause ischemic stroke
IV: instrumental variable
IVW: inverse variance weighted
LAS: large artery atherosclerosis ischemic stroke
LDL: low density lipoprotein cholesterol
MGI: Michigan Genomics Initiative
MR: Mendelian randomization
MR-PRESSO: Mendelian randomization pleiotropy residual sum and outlier test
MVMR: multivariate Mendelian randomization
NTBI: non-transferrin bound iron
SBP: systolic blood pressure
SVS: small vessel ischemic stroke
TC: total cholesterol
TG: triglycerides
TIBC: total iron binding capacity
TSAT: transferrin saturation
UKBB: UK Biobank.

## Acknowledgements

We thank the CARDIoGRAMplusC4D, MEGASTROKE, UK Biobank investigators for making their data publicly available. The MEGASTROKE project received funding from sources specified at http://www.megastroke.org/acknowledgments.html.

## Author’s Contributions

AB designed and conducted the research, analyzed and interpreted the data, and wrote the manuscript; AGC, EKP and KOO interpreted the data and assisted with the manuscript preparation.

## Data and Code Availability

The GWAS summary-level data are publicly available for all exposures and outcomes evaluated. The exposure summary-statistic can be found directly from the original GWAS publication (27). Data on coronary artery disease have been contributed by CARDIoGRAMplusC4D investigators. The UK Biobank, CARDIOGRAMplusC4D consortium, MEGASTROKE, and IL-6 GWAS summary-level data are available in IEU OpenGWAS (https://gwas.mrcieu.ac.uk/). The MR analysis code may be obtained from the corresponding author upon reasonable request.

